# Application of the Robson Classification in Assessing Cesarean Section Rates: An Audit of a Tertiary Care Hospital’s Gynecology Department in Karachi, Pakistan

**DOI:** 10.1101/2023.04.30.23289336

**Authors:** Syed Muhammad Aqeel Abidi, Shahida Abbas, Syeda Tayyaba Fatima Abidi, Namayah Hussain, Sonia Haresh

## Abstract

**Objective:** The increasing rate of cesarean sections (CS) is a significant concern for healthcare providers worldwide. The World Health Organization (WHO) recommends that CS rates should not exceed 10-15% at the population level. The Robson classification system is a widely used tool for assessing and comparing CS rates based on the indications and patient population across different hospitals and obstetric populations. This audit report will help identify areas for improvement in obstetric care practices and facilitate the development of strategies to optimize obstetric care and reduce CS rates, thereby improving maternal and neonatal outcomes.

**Methods:** This retrospective study utilizing hospital records was conducted from January 1st, 2022, to December 31st, 2022, at the Department of Obstetrics and Gynecology in Holy Family Hospital, Karachi, Pakistan. During the study period of one year, a total of 449 cesarean sections were conducted, which were classified into the ten groups of The Robson Classification. Group CS rate, Group size, Relative contribution of group to overall CS rate, and Absolute group contribution to overall CS rate was calculated for each group. Overall indications of CS in all groups was analyzed and based on the results interventions were suggested to be implemented to optimize CS.

**Results:** 1464 women were delivered during this one year period. A total of 507 CS were our ‘population of interest’. Of the cesarean sections performed, 48.8% (247) were elective and 51.2% (260) were emergency. The Robson classification group 5 (51.9%), group 2 (18.5%), and group 10 (14.4%) respectively were the major contributor to the overall CS rate. The main indications for CS were previous history of caesarean section (32.3%), non-progress of labor (21.15%), and cephalopelvic disproportion (16.92%).

**Conclusion:** It is crucial to develop strategies to decrease the unnecessary CS rate while ensuring safe and appropriate obstetric care. Furthermore, there is a need for further research to identify the underlying factors contributing to the high CS rates and to develop effective interventions to address this issue. Implementation of these strategies may lead to improved maternal and fetal outcomes, reduced healthcare costs, and increased patient satisfaction.

## Introduction

The increasing rate of cesarean sections (CS) is a significant concern for healthcare providers worldwide. The World Health Organization (WHO) recommends that CS rates should not exceed 10-15% at the population level, as there is no evidence that higher rates are associated with better maternal or neonatal outcomes (1). However, the rate of CS varies widely across different regions, and in many areas, it is significantly higher than the recommended threshold (2). In 2021 the global rate of CS was 21.1%, Betran, et al. projected that the rate of CS would reach to 30% by 2030 (3).

In Pakistan, the rate of CS has been increasing steadily over the past decade, with one study reporting a rate of 40% between 2017-18 (4). This trend is particularly concerning as the country has a high burden of maternal and neonatal mortality and morbidity, and CS is associated with an increased risk of adverse outcomes for both mothers and infants (5). With the gradual surge in CS rates, the incidences of Morbidly-Adherant Placenta or Placenta accreata also increase, this results in a bad prognosis and is associated with maternal and fetal death (6).

While C-sections can be lifesaving in certain situations, their overuse can lead to increased maternal morbidity and mortality, longer hospital stays, and higher healthcare costs (7). Gibbons et al. estimated in 2010 the global excess CS amounted to USD 2.32 billion (8). Additionally, The incidence of neonatal respiratory complications such as transient tachypnea, surfactant deficiency, and pulmonary hypertension is higher, thus indicating an elevated risk. (9). Therefore, it is important to investigate the factors contributing to the increasing rates of C-sections and to identify effective strategies for reducing their unnecessary use.

To address this issue, healthcare providers must identify the factors contributing to the high rate of CS and develop strategies to reduce unnecessary CS. The Robson classification system is a widely used tool for assessing and comparing cesarean section (CS) rates based on the indications and patient population across different hospitals and obstetric populations. It is a simple and standardized classification system that categorizes all deliveries into ten mutually exclusive groups based on five obstetric characteristics: parity, previous CS, onset of labor, fetal presentation, and gestational age (10). This classification system allows for the identification of groups of women with similar obstetric characteristics and enables comparisons of CS rates within and between groups. The Robson classification system has been adopted by the World Health Organization (WHO) and is recommended as a valuable tool for evaluating obstetric care practices and improving quality of care (1).

In this article, we present the results of an audit of the CS rate at a gynecology department in a tertiary care hospital in Karachi, Pakistan, using the Robson classification system. The significance of this audit lies in its potential to provide valuable insights into obstetric care practices and CS rates at this hospital. This audit report can help clinicians identify areas for improvement in their obstetric care practices and facilitate the development of strategies to optimize obstetric care and reduce CS rates, thereby improving maternal and neonatal outcomes. Furthermore, this audit report can serve as a benchmark for other hospitals in the region and beyond to evaluate their own obstetric care practices and CS rates, ultimately contributing to the improvement of obstetric care on a larger scale.

## Material/Subjects/Patients and methods

### Study design and participants

A retrospective study utilizing hospital records was conducted from January 1st, 2022, to December 31st, 2022, at the Department of Obstetrics and Gynecology in Holy Family Hospital, Karachi, Pakistan. Ethical clearance for the study was granted by the Institute’s Ethical Review Committee.

### Inclusion and exclusion criteria

The study included all patients who underwent cesarean delivery during the designated period, while all vaginal deliveries were excluded from the analysis.

### Study setting

Holy Family Hospital, Karachi, Pakistan is a tertiary care hospital with an obstetrics and gynecology department with over 1000 normal deliveries per year. Holy Family Hospital is a teaching institute with the obstetrics and gynecology department being recognized as a postgraduate training program by College of Physicians and Surgeons Pakistan (CPSP).

### Study procedure

During the study period of one year, a total of 449 cesarean deliveries were conducted at the department of Obstetrics and Gynecology, Holy Family Hospital, Karachi, Pakistan. These deliveries were classified into the ten groups of The Robson Classification, as presented in Figure 1. The hospital records of the medical record section were reviewed to extract relevant information for each case. The total number of deliveries was matched with the labor room record. A standardized proforma was designed and utilized to record various details of each cesarean section case, including maternal characteristics (age, parity, mode of previous deliveries, gestational age, number of fetuses, fetal presentation, previous CS and its indications, and onset of labor), fetal outcomes (birth weight, APGAR score, and fetal complications), and maternal outcomes (postpartum hemorrhage, anemia, wound infection, need for blood transfusion, ruptured uterus, ICU admission, and maternal mortality). All the data were collected and analyzed by the research team in accordance with the guidelines of the Robson classification system. The study adhered to the principles of data confidentiality and patient privacy.

### Attach Figure 1

**Figure 1:**
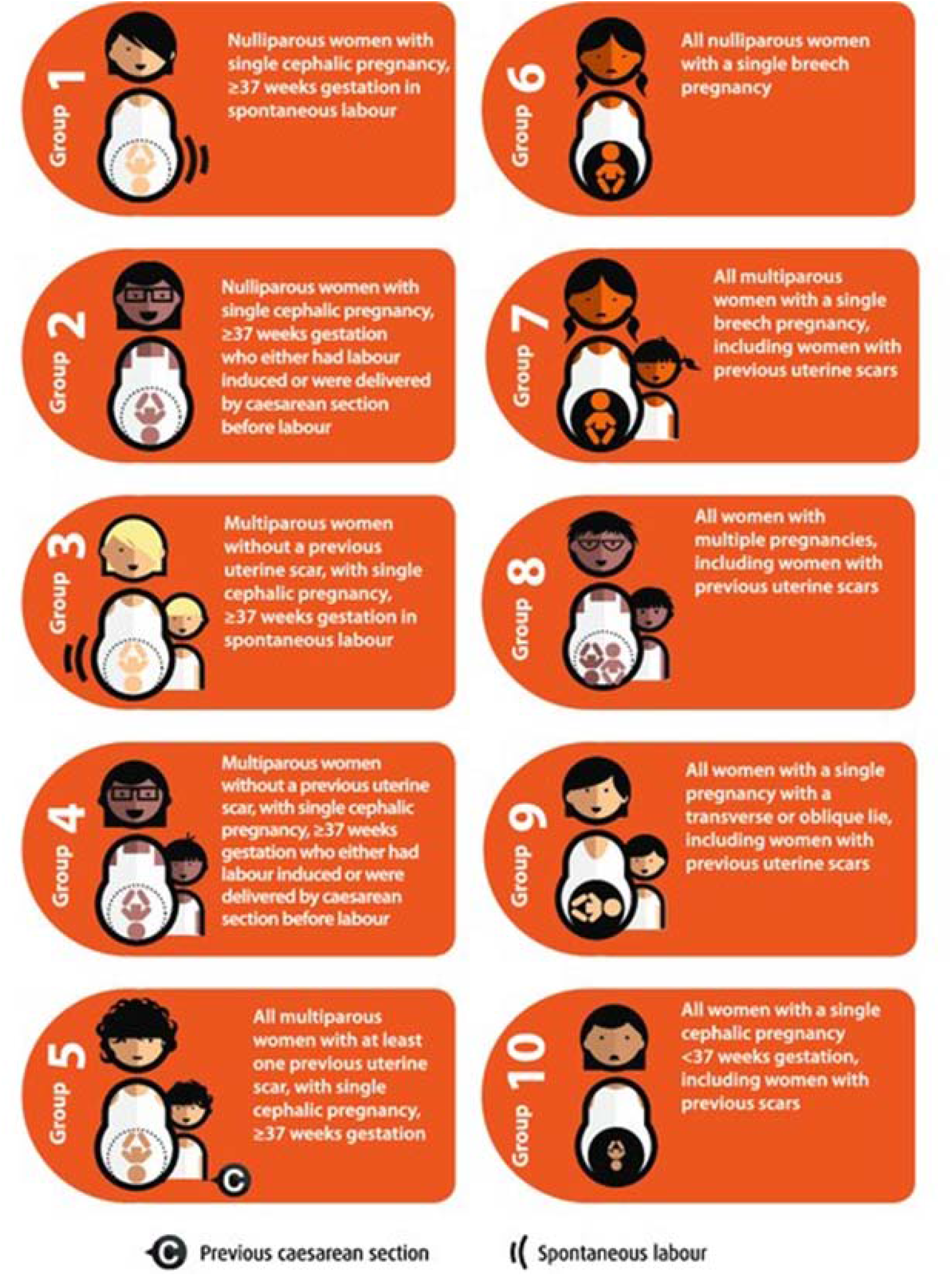
Robson’s Classification.

### The Robsons Classification

The Robson classification system is a tool that enables the evaluation of the obstetric patient population and the Cesarean section (CS) rates in a standardized and comparable manner across different settings. This system categorizes all deliveries into ten mutually exclusive groups based on five key obstetric characteristics: parity, onset of labor, gestational age, fetal presentation, and number of fetuses (Figure 1). These groups can be used to identify areas of overuse or underuse of CS, and to monitor changes in CS rates over time. In our study, we applied the Robson classification system to the study population of the gynecology department at a tertiary care hospital in Karachi, Pakistan to assess the CS rates and to identify potential areas for improvement in clinical practice.

### Statistical analysis

The data from individual cases were recorded in a Microsoft Excel spreadsheet, and statistical analysis was performed using SPSS software, version 25. Categorical variables were expressed as percentages. Descriptive statistics were used to summarize the demographic and obstetric characteristics of the study population. The size of each Robson group, CS rate, and their respective contribution to the overall CS rate were determined. Indications for CS in each group were analyzed. The CS rates were then compared across the different Robson groups to identify the groups with the highest CS rates. Statistical significance was assessed using chi-square tests with a p-value of less than 0.05 indicating significance. Odds ratios (OR) and 95% confidence intervals (CI) were also calculated to examine the association between Robson groups and CS rates. Multivariable logistic regression analysis was performed to adjust for potential confounding factors.

## Results

During 1^st^ January 2022 and 31^st^ December 2022, 1464 women met the inclusion criteria and were included in the study. Month-wise number of caesarean sections are given in Figure 1. A total of 507 deliveries were included in the study, which was our ‘population of interest’. A total of 967 were Spontaneous Vaginal Deliveries (SVDs) and were excluded from the study. 67.06% (340) of the patients belonged to the main city, whereas 32.94% (167) belonged to the peripheral areas. This yielded a CS rate of 54.2%. Of the cesarean sections performed, 48.8% (247) were elective and 51.2% (260) were emergency.

**Figure 2:**
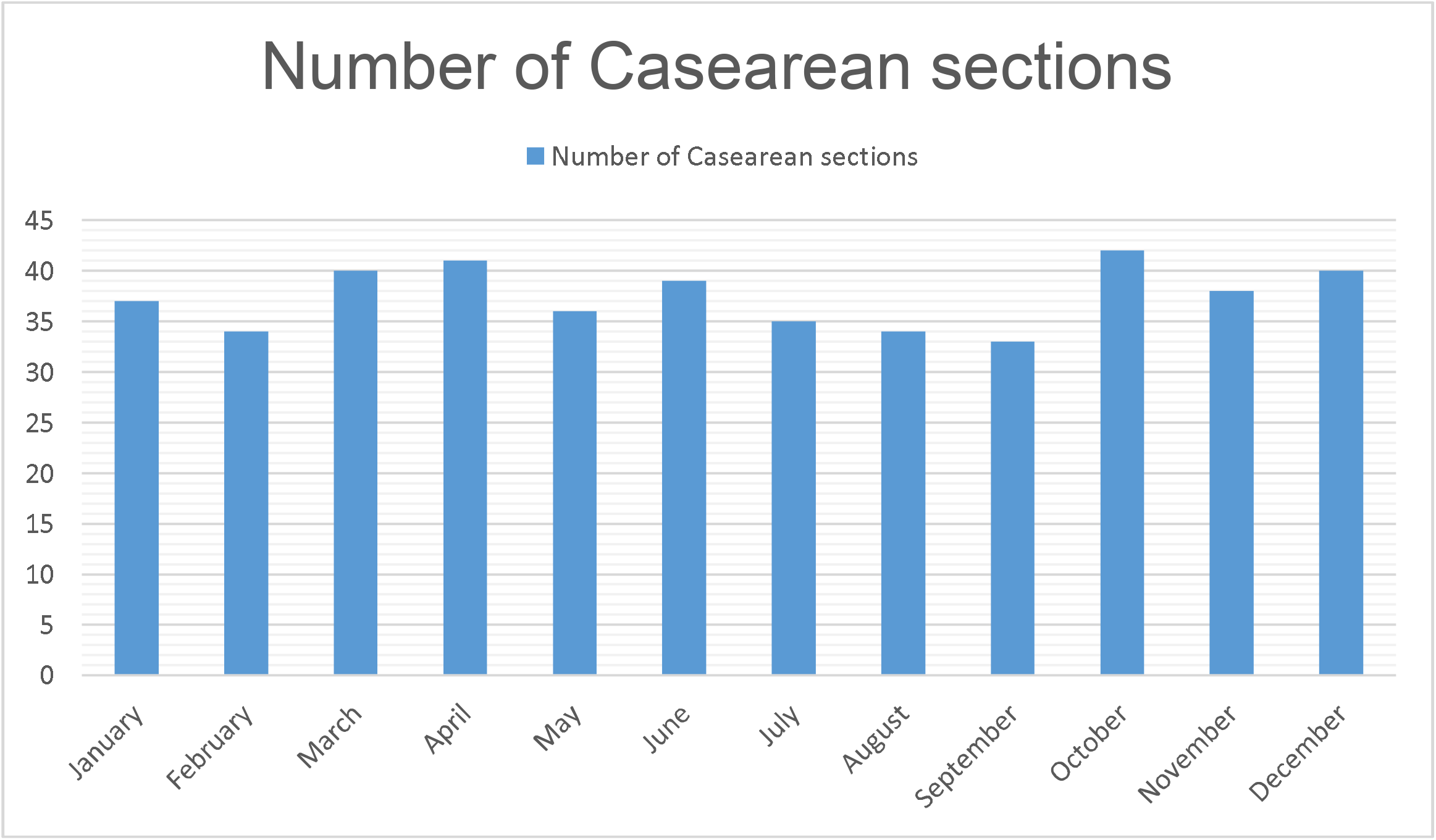
Month-wise distribution of CS.

Most of the study participants (323) were in the age group of 21-30 years (63.7%). Nulliparous women accounted for 37.3% (188) of the study population, while multiparous women accounted for 62.7%. Women with a history of previous CS constituted 64.09% (323) of the study participants. The majority of the CS (99%) were performed at term between 37 and 40 weeks of gestation.

**Figure 3:**
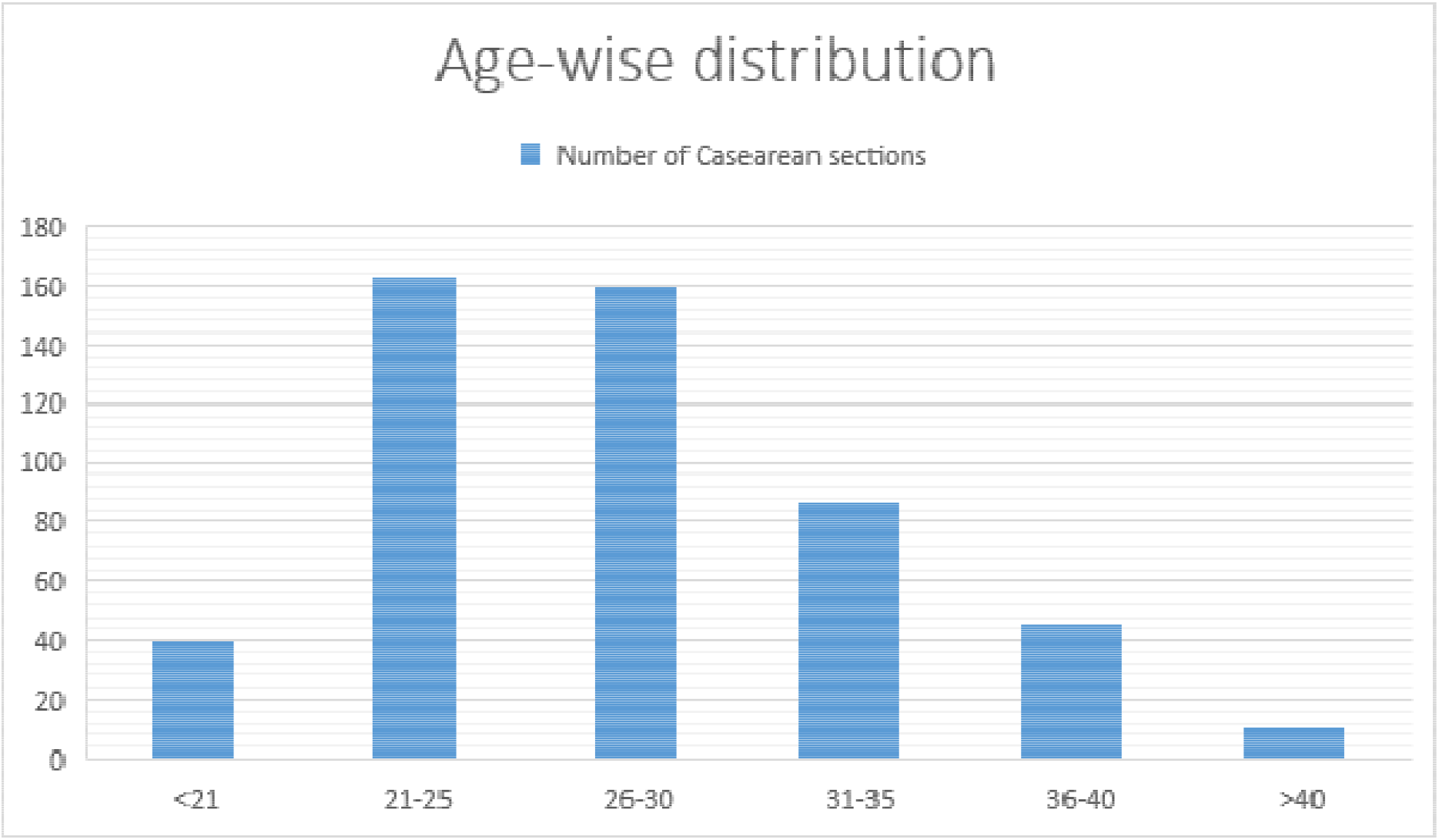
Age-wise distribution of CS.

The Robson classification showed that group 5 (Previous cesarean section, singleton, cephalic, ≥□37 weeks’ gestation) was the highest contributor to the overall CS rate, contributing 51.9% of all CS and 27.5% in ratio to all deliveries at hospital. Group 2 (Nulliparous, single cephalic, ≥□37 weeks, induced or CS before labor) was the second highest contributor, contributing 18.5% to the overall CS and 9.82% in ratio to all deliveries at hospital. The third highest contributors were group 10 (All singleton, cephalic, <□37 weeks’ gestation pregnancies (including previous cesarean section)) contributing 14.4% to the overall CS rate and 7.63% in ratio to all deliveries at hospital. The least contributor to the overall CS rate was group 9 (All women with a single pregnancy in transverse or oblique lie (including those with previous cesarean section)), contributing 0.39% of all CS and 0.21% in ratio to all deliveries at hospital. The remaining groups (groups 1, 3, 4, 6, 7, and 8) contributed 15.2% of all CS and 9.05% in ratio to all deliveries at hospital.

The main indications for CS in the 260 patients who underwent emergency caesarean sections were previous history of caesarean section (32.3%), non-progress of labor (21.15%), cephalopelvic disproportion (16.92%), fetal distress (8.46%), breech presentation (6.54%), pre-eclampsia (6.15%), refusal for VBAC (5.00%), low-lying placenta (3.09), and transverse lie (0.39%).

## Discussion

The present audit report assessed the cesarean section (CS) rates using the Robson Classification at a tertiary care hospital’s gynecology department in Karachi, Pakistan. The Robson’s classification, designed in 2001, is often referred to as the ‘Ten Group Classification System (TGCS)’, as it divides Caesarean Sections into ten groups based on the gestational age of the pregnancy, several pregnancy-related characteristics (parity, position, presentation, etc.), prior obstetrical history, and labour and delivery procedures and allows identification of the groups that contributed the most to the overall CS (10). It was officially endorsed by WHO in 2015 as the gold standard to monitor Caesarean Sections (11).

Analyzing the Cesarean Section (CS) rate is crucial in determining the quality of maternal healthcare and ensuring the safety of both the mother and the child during childbirth. A high CS rate may suggest overuse of the technique, which might put the mother and infant at needless danger and complicate matters (12). On the other hand, a Low CS rates may imply underuse of the operation, which, particularly in some patient populations, may lead to avoidable maternal and newborn morbidity and death (13). Therefore, analyzing the CS rate is an essential tool for evaluating the effectiveness and efficiency of maternal healthcare systems, identifying areas for improvement, and ensuring that women receive appropriate and timely care during childbirth. The objective of this study is to determine the CS rate and investigate the factors contributing to it in the tertiary care hospital and compare it with hospitals in similar setting, with the aim of identifying strategies to improve the quality and safety of maternal healthcare.

The findings indicate a CS rate of 54.2%, which is higher than the World Health Organization’s recommended rate of 10-15% (1). The highest contributor was group 5, which consisted of women with previous cesarean section, singleton, cephalic, ≥□37 weeks’ gestation. This finding is consistent with other studies (2), group 5 was the second highest contributor in other studies (14, 15). Majority of the sections happened due to history of previous caesarean sections, which was similar to other studies (14-16). A commonality in all these studies including ours (14-16) was the low rate or absence of Caesarean section due to refusal for Vaginal Birth After Cesarean (VBAC), this indicates that the doctors in developing region are not promoting the use of VBAC. There is a need to investigate the causes of doctors not offering VBAC option to patients but possible reasons might include the recent literature showing decreased success rates of VBAC (17) and increased risk of legal proceedings over failure of VBAC (18).

Previous history of CS being the major indication for CS highlights the importance of promoting vaginal birth after cesarean (VBAC) in eligible women to reduce the rate of repeat CS. The American College of Obstetricians and Gynecologists recommends VBAC as a safe and appropriate option for most women with a previous CS (19).

The second highest contributor was group 2, which consisted of nulliparous, single cephalic, ≥□37 weeks who were either induced or had CS before labor. This finding is consistent with the findings by Ansari A et al. at a tertiary care hospital in Pakistan (16), but the other studies didn’t have group 2 as major contributor (14, 15). This is troubling since nulliparous women having a high likelihood of CS, can result in needless risks such greater maternal morbidity and death, lengthier hospital stays, and higher healthcare expenses (20). Therefore, it is crucial to implement policies to reduce the rate of CS in this group.

The percentage of elective Caesarean Sections was very high, this indicates either the community is ill-informed about Caesarean sections and its implications or that they aren’t being counseled well by the admitting doctors. Hence, there is a need to conduct an awareness campaign about the benefits and pitfalls of caesarean sections. The junior doctors should be trained to conduct the counseling of the expecting mothers and be well-versed in indications for caesarean sections.

The findings of this study suggest that there is a need for strategies to optimize the use of CS and reduce the overall CS rate in the hospital’s gynecology department. One approach could be to improve labor management, including the appropriate use of induction of labor and fetal monitoring, to reduce the rate of non-progress of labor and fetal distress, which were among the top indications for CS in this study. Additionally, increasing access to VBAC and implementing VBAC-friendly policies may help to reduce the rate of repeat CS. These strategies should be implemented with caution and should take into account the individual needs and preferences of women and their families.

While, the study utilized a standardized protocol for data collection and a validated classification system (The Robson Classification) to evaluate the CS rates in a comparable manner across different settings and had a large sample size of 449 cesarean deliveries in one year, which provides a good representation of the population in the study setting. However, this study was conducted in only one hospital in Karachi, Pakistan, which limits the generalizability of the findings to other settings. The study did not include vaginal deliveries in the analysis, which may have provided a more comprehensive picture of the overall obstetric population. The study was retrospective in nature, which may lead to incomplete or missing data, which limits the ability to control for confounding variables. The study did not record information about the reasons for the high CS rates individually in each Robson groups, which limits the ability to identify specific areas for improvement in clinical practice. The study did not collect information on some potentially important variables, such as BMI, and fetal weight, which may have influenced the CS rate.

In the future, we will be designing strategies based on the above given results and post-implementation we will repeat this audit to look at the results after the implementation, which will indicate the success of the interventions and give directions for future interventions.

## Conclusion

The analysis of Cesarean section (CS) rate revealed that the CS rate was more than 3-folds higher than the recommended threshold set by the WHO. Primary indication for CS was previous history of CS. It is crucial to develop strategies to decrease the unnecessary CS rate while ensuring safe and appropriate obstetric care. Furthermore, there is a need for further research to identify the underlying factors contributing to the high CS rates and to develop effective interventions to address this issue. The findings suggest the importance of promoting VBAC and implementing strategies to reduce the rate of non-progress of labor and fetal distress to lower the overall CS rate. The implementation of these strategies may lead to improved maternal and fetal outcomes, reduced healthcare costs, and increased patient satisfaction.

## Data Availability

All data produced in the present work are contained in the manuscript

## Acknowledgement

There were no acknowledgments to make for this research project.

## Disclaimer

The opinions expressed in this paper are those of the authors and do not necessarily reflect the views of any organization or institution with which they may be affiliated. The information presented in this paper is intended for academic and research purposes only and should not be construed as professional advice or recommendations.

## Conflict of interest

The authors declare no conflict of interest related to this research project. They have no financial or personal relationships that could influence the results or interpretation of this study.

## Funding disclosure

The authors declare that no funding was received for this research project. This paper is the result of an independent research effort by the authors and any opinions or conclusions presented in this study are solely those of the authors.

The authors declare that data supporting the findings of this study are available within the article. The article is the author(s) original work

The article has not received prior publication and is not under consideration for publication elsewhere All the authors have seen and approved the manuscript being submitted

The author(s) abide by the copyright terms and conditions of publishing journal

---

**Table 1:**
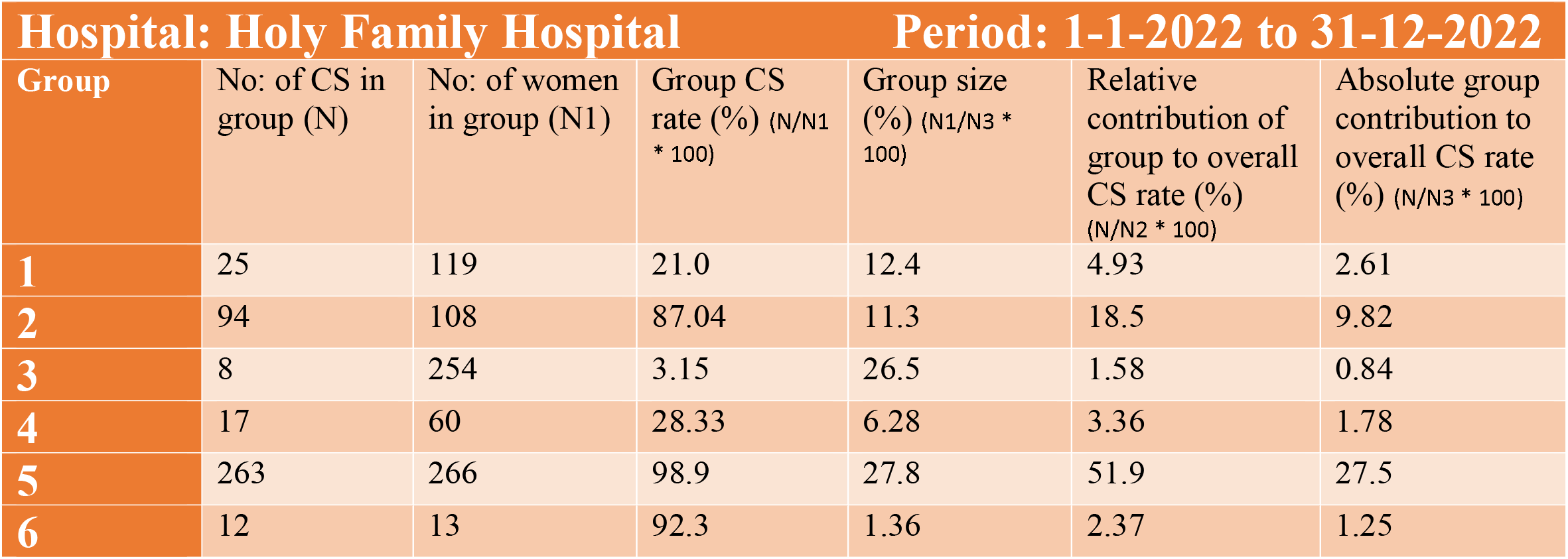
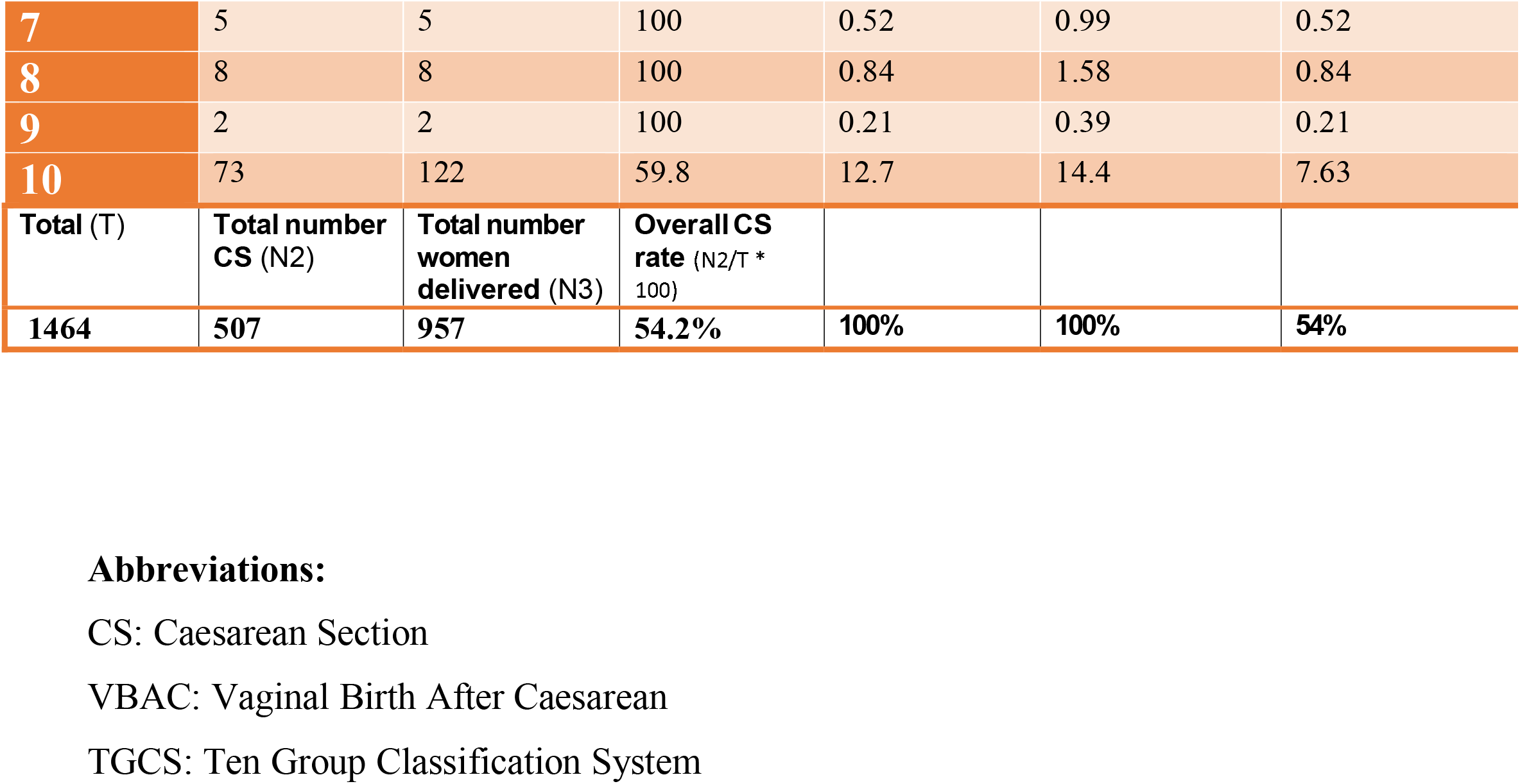
Distribution of CS by different subgroups of TGCS.

| Hospital: Holy Family Hospital |  |  |  | Period: 1-1-2022 to 31-12-2022 |  |  |
| --- | --- | --- | --- | --- | --- | --- |
| Group | No: of CS in group (N) | No: of women in group (N1) | Group CS rate (%) (N/N1 * 100) | Group size (%) (N1/N3 * 100) | Relative contribution of group to overall CS rate (%) (N/N2 * 100) | Absolute group contribution to overall CS rate (%) (N/N3 * 100) |
| 1 | 25 | 119 | 21.0 | 12.4 | 4.93 | 2.61 |
| 2 | 94 | 108 | 87.04 | 11.3 | 18.5 | 9.82 |
| 3 | 8 | 254 | 3.15 | 26.5 | 1.58 | 0.84 |
| 4 | 17 | 60 | 28.33 | 6.28 | 3.36 | 1.78 |
| 5 | 263 | 266 | 98.9 | 27.8 | 51.9 | 27.5 |
| 6 | 12 | 13 | 92.3 | 1.36 | 2.37 | 1.25 |
| 7 | 5 | 5 | 100 | 0.52 | 0.99 | 0.52 |
| 8 | 8 | 8 | 100 | 0.84 | 1.58 | 0.84 |
| 9 | 2 | 2 | 100 | 0.21 | 0.39 | 0.21 |
| 10 | 73 | 122 | 59.8 | 12.7 | 14.4 | 7.63 |
| Total (T) | Total number CS (N2) | Total number women delivered (N3) | Overall CS rate (N2/T * 100) |  |  |  |
| 1464 | 507 | 957 | 54.2% | 100% | 100% | 54% |

**Abbreviations:**
CS: Caesarean Section
VBAC: Vaginal Birth After Caesarean
TGCS: Ten Group Classification System

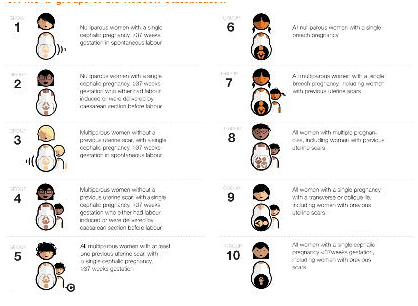

## Notes

### Competing Interest Statement

The authors have declared no competing interest.

### Funding Statement

This study did not receive any funding

### Author Declarations

Holy Family Hospital Ethical Review Board, clearance was granted

